# Resampling Methods for Class Imbalance in Clinical Prediction Models: A Systematic Review and Meta-Regression Protocol

**DOI:** 10.1101/2025.05.19.25327868

**Authors:** Osama Abdelhay, Adam Shatnawi, Hassan Najadat, Taghreed Altamimi

**Affiliations:** Department of Data Science and Artificial Intelligence, Princess Sumaya University for Technology, Amman, Jordan; Department of Computer Science, Jordan University of Science and Technology, Irbid, Jordan; College of Engineering and Advanced Computing, Alfaisal University, Riyadh, Saudi Arabia

## Abstract

**Introduction:** Class imbalance—situations where clinically important “positive” cases form <30 % of the dataset—systematically degrades the sensitivity and fairness of medical prediction models. Although data-level techniques such as random oversampling, random undersampling and SMOTE, and algorithm-level approaches like cost-sensitive learning, are widely used, the empirical evidence describing when these corrections improve model performance remains fragmented across diseases and modelling frameworks. This protocol outlines a scoping systematic review with meta-regression that will map and quantitatively summarise 15 years of research on resampling strategies in imbalanced clinical datasets, addressing a critical methodological gap in trustworthy medical AI.

**Methods and analysis:** We will search MEDLINE, EMBASE, Scopus, Web of Science Core Collection and IEEE Xplore, plus grey-literature sources (medRxiv, arXiv, bioRxiv) for primary studies (2009 – 31 Dec 2024) that apply at least one resampling or cost-sensitive method to binary clinical prediction tasks with a minority-class prevalence <30 %. No language restrictions will be applied. Two reviewers will screen records, extract data with a piloted form and document the process in a PRISMA flow diagram. A descriptive synthesis will catalogue clinical domain, sample size, imbalance ratio, resampling technique, model type and performance metrics where≥10 studies report compatible AUCs, a random-effects mixed-effects meta-regression (logit-transformed AUC) will examine moderators including imbalance ratio, resampling class, model family and sample size. Small-study effects will be probed with funnel plots, Egger’s test, trim-and-fill and weight-function models; influence diagnostics and leave-one-out analyses will assess robustness. Because this is a methodological review, formal clinical risk-of-bias tools are optional; instead, design-level screening, influence diagnostics and sensitivity analyses will ensure transparency.

**Discussion:** By combining a broad conceptual map with quantitative estimates, this review will establish when data-level versus algorithm-level balancing yields genuine improvements in discrimination, calibration and cost-sensitive metrics across diverse medical domains. The findings will guide researchers in choosing parsimonious, evidence-based imbalance corrections, inform journal and regulatory reporting standards, and highlight research gaps, such as the under-reporting of calibration and misclassification costs, that must be addressed before balanced models can be trusted in clinical practice.

**Systematic review registration:** INPLASY202550026

## Introduction

Medical prediction datasets are frequently imbalanced, with the clinically important “positive” class representing fewer than 30 % of observations. Such skew systematically biases classical (e.g., logistic regression) and modern machine-learning classifiers toward the majority class, eroding sensitivity for the minority group [1–3].

To mitigate this threat, a family of data-level resampling techniques—random oversampling (ROS), random undersampling (RUS), and the Synthetic Minority Oversampling Technique (SMOTE) adjust the training data before modelling [1,4,5]. While widely adopted, ROS can overfit duplicates, RUS discards potentially informative cases, and SMOTE or its variants may inject unrealistic synthetic examples [6–9].

Evidence comparing resampling to alternative strategies remains inconclusive. An extensive systematic review showed no consistent performance advantage of machine learning over logistic regression when event-per-variable ratios were adequate [10].

Moreover, simulation and empirical studies demonstrate that adequate sample size planning, rather than aggressive post-hoc balancing, often eliminates the need for resampling [11–16].

At the algorithm level, cost-sensitive learning penalises minority-class errors directly and can outperform data-level methods, yet it is rarely reported in medical AI research [4,17].

Developments in binary classification theory—from early statistical formulations to perceptrons, support vector machines, and boosted ensembles—underline that model choice interacts with class distribution and cost structure [18–21].

Against this backdrop, we will conduct a scoping systematic review with meta-regression to (i) map resampling and cost-sensitive strategies used in imbalanced medical datasets, (ii) quantify their impact on discrimination and calibration, and (iii) identify methodological moderators and research gaps. This protocol describes the planned methods.

### Objectives Primary objective

To determine, across clinical prediction studies with binary outcomes and minority-class prevalence < 30 %, whether applying data-level resampling or algorithm-level cost-sensitive strategies measurably improves model performance relative to training on the original imbalanced data.

### Specific objectives

1. **Evidence mapping** – Catalogue the full range of imbalance-correction strategies (oversampling, undersampling, hybrids, weighted or focal-loss models) reported between 2009 and 2024, together with the clinical domains, dataset sizes, imbalance ratios, and modelling frameworks in which they are used.
2. **Comparative effectiveness** – Quantify and compare discrimination (e.g., AUC, sensitivity, specificity) and, where available, calibration metrics achieved by
  ∘ oversampling,
  ∘ undersampling,
  ∘ hybrid pipelines, and
  ∘ cost-sensitive algorithms, against models trained without any balancing.
3. **Moderator analysis** – Using mixed-effects meta-regression, evaluate how study-level characteristics (imbalance ratio, sample size, number of predictors, model family, and clinical domain) modify the effectiveness of each imbalance-correction strategy.
4. **Bias and robustness assessment** – Probe small-study effects, publication bias, and influential outliers through funnel-plot diagnostics, trim-and-fill, weight-function models, and leave-one-out analyses; gauge how these factors affect pooled estimates.
5. **Methodological gap identification** – Highlight recurrent pitfalls, such as neglecting calibration, misclassification costs, or external validation, and formulate evidence-based recommendations for future research and reporting.

## Methods

This protocol adheres to the PRISMA-P [22] and PRISMA-ScR [23] guidelines and has been registered with INPLASY (ID INPLASY202550026) (S1 File). Any amendments will be documented in the INPLASY record.

### Eligibility criteria (PICOTS)

- **Population:** Clinical prediction studies that analyse binary outcomes with an explicit minority-class prevalence < 30 %. For this review, a *binary outcome* is restricted to diagnostic, prognostic, or treatment-response predictions in which the dependent variable has exactly two mutually exclusive states (e.g., disease present/absent).
- **Interventions:** Data-level resampling (random oversampling, random undersampling, SMOTE or variants, hybrid pipelines) and algorithm-level cost-sensitive techniques (weighted losses, focal loss).
- **Comparators:** Models trained on the original imbalanced data and/or alternative resampling or weighting strategies.
- **Outcomes:** Primary—AUC; secondary—sensitivity, F1-score, specificity, balanced accuracy, calibration metrics, and reported mis-classification costs.
- **Timing:** Publications from 1 Jan 2009 to 31 Dec 2024.
- **Study design:** Retrospective or prospective primary studies (including model-development and validation papers) and systematic reviews that re-analyse primary data. Simulation-only papers, non-binary tasks, or abstracts lacking methods are excluded. Studies focused exclusively on radiomics, image-segmentation pipelines, or pixel-level classification tasks will be excluded because these do not output patient-level binary predictions.

### Information sources and search strategy

Searches were executed in MEDLINE (PubMed), EMBASE, Scopus, Web of Science Core Collection and IEEE Xplore. Grey-literature repositories (medRxiv, arXiv, bioRxiv) and code platforms (GitHub) were also screened. A peer-reviewed strategy combined controlled vocabulary and free-text terms for *class imbalance, resampling*, and *clinical prediction*; an example MEDLINE string is provided in S2 File. No language limits were applied, but non-English full texts had to be translatable.

### Study selection

Search results will be imported into Zotero for deduplication [24] and prioritised with ASReview [25]. Two reviewers will independently screen titles/abstracts, followed by full texts, resolving conflicts by consensus or third adjudication. Reasons for exclusion will be recorded and displayed in a PRISMA flow diagram [26]. Data missing from the full text will be requested from authors (two-week window).

### Data extraction

A piloted, standardised form will capture bibliometrics, clinical domain, sample size, imbalance ratio, resampling strategy, model family, performance metrics, calibration statistics, and cost-sensitive measures. Two independent reviewers will extract all items in duplicate into a REDCap (v14.0.19) database. A third reviewer will run the built-in comparison report, reconcile discrepancies, and export a single verified dataset.

Statistical analyses will be performed in R (v4.4.0) using the *metafor* package (v4.8-0), *dplyr* (v1.1.4) and *ggplot2* (v3.5.2). All code and a session-info file will be deposited in the OSF repository on publication.

### Outcomes and effect measures

The primary effect size will be the logit-transformed AUC. When compatible statistics are reported, we will also collect sensitivity, F1 Score, calibration slope, Brier score, and cost-based metrics. External validation results will be catalogued separately.

### Risk-of-bias and methodological quality

Because the review focuses on methodological interventions rather than clinical effects, formal study-level tools (e.g., PROBAST) will be optional [27]. Instead, we will apply design-level screening for reproducibility, influence diagnostics (Cook’s distance [28], studentised residuals [31]), and small-study-effect tests (funnel plot [31], Egger’s regression [29], Vevea–Hedges weight-function [30]) to inform sensitivity analyses. We will still chart whether studies report blinding, missing-data handling, and external validation; we plan to slot these parts into a supplementary risk-of-bias table.

### Data synthesis

*Phase 1—Descriptive mapping: Tables and visualisations (e.g., heat maps, temporal plots) will summarise trends in resampling use, model type, imbalance severity, and performance*.

*Phase 2 — Quantitative synthesis*: random-effects meta-regression of logit-AUC will examine moderators (imbalance ratio, sample size, resampling class, model family). REML estimator and Knapp-Hartung confidence intervals will be employed [31].

Heterogeneity will be assessed with τ^2^ and I^2^ [31]; leave-one-out analyses will test robustness. Analyses will be implemented in R (metafor, dplyr, ggplot2) [31].

### Subgroup and sensitivity analyses

Planned subgroup contrasts include oversampling vs undersampling, hybrid vs single-technique pipelines, cost-sensitive vs data-level only, high (>20 %) vs very-low (<5 %) minority prevalence, and deep-learning vs traditional models. Sensitivity analyses will exclude high-influence studies, studies without external validation, and those lacking calibration reporting. Imbalance ratio (IR) will be stratified a priori into four bins: very rare < 5 %, rare 5–10 %, moderate 10–20 %, and mild 20–30 % [6]. If any bin has < 10 studies, it will be merged with the next wider bin. For meta-regression, these bins will be dummy-coded (reference = mild), and IR will also be modelled as a restricted cubic spline to test linearity. Sensitivity analyses will repeat the model using two dichotomies (< 10 % vs ≥ 10 %; < 20 % vs ≥ 20 %).

### Living review plan

Given rapid methodological advances, automated database alerts will rerun the search annually; new eligible studies will be screened and, where appropriate, integrated into updated meta-analyses, with version history transparently logged.

## Discussion

Class imbalance remains one of the most stubborn threats to safe clinical prediction: skewed data encourage algorithms to optimise *overall* accuracy at the expense of rare—but clinically essential—events. Algorithm-level approaches that embed explicit mis-classification penalties can theoretically offset this bias [32,33]. At the same time, recent deep-learning innovations such as deep belief nets and focal-loss functions promise further gains in high-dimensional settings [34,35]. Yet the empirical value of these strategies has never been synthesised systematically across the medical spectrum. Our planned scoping review with meta-regression addresses a critical methodological gap.

### Anticipated challenges

- **Extreme heterogeneity**: preliminary scoping suggests wide dispersion in clinical domains, imbalance ratios, sample sizes, and metrics. Even when studies report AUC, transformation to a common logit scale may not fully harmonise differences in test–set construction and cross-validation folds.
- **Inconsistent reporting**: fewer than one in ten studies publish calibration indices, and details of cost-sensitive losses are often relegated to supplementary code or omitted entirely.
- **Sparse external validation**: most papers evaluate performance on random internal splits; true generalisability remains unknown.
- **Publication and small-study effects:** funnel-plot asymmetry is expected because smaller datasets often adopt aggressive oversampling that inflates apparent discrimination.
- **Metric multiplicity**: sensitivity, specificity, F-score, precision-recall AUC and balanced accuracy are reported idiosyncratically, complicating quantitative synthesis.

### Strengths

- **Breadth of evidence**: five bibliographic databases plus grey-literature repositories capture 15 years of work, producing the most extensive curated corpus of imbalance-related prediction studies.
- **Dual synthesis**: A descriptive map is paired with a random-effects meta-regression that tests moderators such as imbalance severity, sample size, and model family, offering granular insight unavailable in narrative reviews.
- **Rigorous bias diagnostics**: influence statistics, funnel-plot tests, trim-and-fill, and Vevea–Hedges models will quantify the robustness of pooled estimates, mitigating the optimism plaguing model-development literature.
- **Technology-enabled workflows**: machine learning–assisted screening via ASReview accelerates and transparently documents selection decisions [25].
- **Alignment with contemporary guidance**: search, extraction, and reporting follow PRISMA 2020 extensions to enhance reproducibility and uptake [26].

## Limitations

Despite these safeguards, several constraints remain. First, residual heterogeneity is inevitable: even a comprehensive meta-regression may explain only a modest fraction of between-study variance. Second, the decision to use AUC as the primary effect size risks overlooking threshold-dependent performance and real-world utilities. Third, cost-sensitive studies may still be too few or inconsistently reported to support quantitative pooling, forcing a descriptive treatment that limits formal comparisons with resampling. Fourth, living-review updates will depend on the speed at which newly published work reports compatible statistics—the review could lag very recent methodological advances.

### Potential impact and influence on practice

By establishing when and for whom resampling or weighting truly adds value, the review will help data scientists avoid reflexive oversampling that can obscure calibration or foster over-fitting. Evidence that cost-sensitive losses rival data-level balancing could shift practice toward simpler, loss-function–centric pipelines already available in mainstream frameworks [32–35]. Clinicians and journal editors could use the findings to demand fuller reporting of calibration, confusion matrices and mis-classification costs, accelerating the adoption of emerging AI reporting extensions (e.g., TRIPOD-AI), see also [26]. Regulators may likewise reference our recommendations when assessing the fairness of deployed diagnostic or prognostic models.

### Future directions

The mapped gaps suggest four priorities:

- **Prospective, multi-centre cohorts with rare outcomes** to test whether cost-sensitive and focal-loss networks outperform oversampling in truly out-of-sample settings.
- **Standardised reporting templates** that mandate disclosure of class distribution, sampling strategy, calibration and decision-curve analysis; our findings can feed directly into upcoming guideline revisions.
- **Generative augmentation and domain-adapted GANs**: Early evidence (e.g., synthetic EEG and radiology data) hints at privacy-preserving promise but requires rigorous external validation [36].
- **Continuous evidence surveillance** through annual database alerts and semi-automated screening pipelines aligns with the living-review paradigm and ensures the conclusions remain current as new imbalance-handling techniques emerge [25,26].

The planned review will quantify the performance lift (or degradation) attributable to balancing strategies and frame a research agenda aimed at more reproducible, cost-aware and clinically grounded predictive modelling.

## Data Availability

No datasets were generated or analysed during the current study. All relevant data from this study will be made available upon study completion.

