## Supplementary material for "Resampling Methods for Class Imbalance in Clinical Prediction Models: A Systematic Review and Meta-Regression Protocol": S1

**Identification of studies via databases and registers**

Records removed *before screening*:

Duplicate records removed (n = )

Records marked as ineligible by automation tools (n = )

Records removed for other reasons (n = )

Records identified from*:

Databases (n = )

Registers (n = )

**Identification**

Records screened

(n = )

Records excluded**

(n = )

Reports sought for retrieval

(n = )

Reports not retrieved

(n = )

**Screening**

Reports assessed for eligibility

(n = )

Reports excluded:

Reason 1 (n = )

Reason 2 (n = )

Reason 3 (n = )

etc.

Studies included in review

(n = )

Reports of included studies

(n = )

**Included**

*Consider, if feasible to do so, reporting the number of records identified from each database or register searched (rather than the total number across all databases/registers).

**If automation tools were used, indicate how many records were excluded by a human and how many were excluded by automation tools.

Source: Page MJ, et al. BMJ 2021;372:n71. doi: 10.1136/bmj.n71.

This work is licensed under CC BY 4.0. To view a copy of this license, visit <https://creativecommons.org/licenses/by/4.0/>
