## Supplementary material for "Resampling Methods for Class Imbalance in Clinical Prediction Models: A Systematic Review and Meta-Regression Protocol": S2

Table A1: Ready-to-paste search queries with limit (2009 to 30^th^ of April 2024)

| Database | Syntax-ready string* |
| --- | --- |
| PubMed / MEDLINE | ((imbalanc*[tiab] OR "minority class"[tiab] OR "skewed dataset*"[tiab]) AND (oversampl*[tiab] OR undersampl*[tiab] OR SMOTE[tiab] OR "synthetic data"[tiab] OR "cost‑sensitive"[tiab] OR weighting[tiab]) AND (clinic*[tiab] OR medic*[tiab] OR health*[tiab] OR patient*[tiab] OR diagnosis[tiab] OR prognos*[tiab]) AND (predict*[tiab] OR classification[tiab] OR "logistic regression"[tiab] OR "machine learning"[tiab] OR "artificial intelligence"[tiab])) AND ("2009/01/01"[dp] : "2024/12/30"[dp]) [PubMed](https://pubmed.ncbi.nlm.nih.gov/help/?utm_source=chatgpt.com) |
| EMBASE | (imbalanc* OR "minority class*" OR "skewed dataset*").ti,ab,kw. AND (oversampl* OR undersampl* OR SMOTE OR "synthetic data" OR cost‑sensitive OR weighting).ti,ab,kw. AND (clinic* OR medic* OR health* OR patient* OR diagnosis OR prognos*).ti,ab,kw. AND (predict* OR classification OR "logistic regression" OR "machine learning" OR "artificial intelligence").ti,ab,kw. ↲ limit 1 to yr="2009 - 2024" [browse.welch.jhmi.edu](https://browse.welch.jhmi.edu/searching/embase-search-tips?utm_source=chatgpt.com) |
| Scopus | TITLE-ABS-KEY ( imbalanc* OR "minority class" OR "skewed dataset*" ) AND TITLE-ABS-KEY ( oversampl* OR undersampl* OR SMOTE OR "synthetic data" OR "cost‑sensitive" OR weighting ) AND TITLE-ABS-KEY ( clinic* OR medic* OR health* OR patient* OR diagnosis OR prognos* ) AND TITLE-ABS-KEY ( predict* OR classification OR "logistic regression" OR "machine learning" OR "artificial intelligence" ) AND PUBYEAR > 2008 AND PUBYEAR < 2025 [Stack Overflow](https://stackoverflow.com/questions/35526682/scopus-search-title-abs-key/42181577?utm_source=chatgpt.com) |
| Web of Science Core Collection | TS=(imbalanc* OR "minority class" OR "skewed dataset*") AND TS=(oversampl* OR undersampl* OR SMOTE OR "synthetic data" OR "cost-sensitive" OR weighting) AND TS=(clinic* OR medic* OR health* OR patient* OR diagnosis OR prognos*) AND TS=(predict* OR classification OR "logistic regression" OR "machine learning" OR "artificial intelligence") AND PY=(2009-2024) [images.webofknowledge.com](https://images.webofknowledge.com/images/help/WOS/hs_advanced_fieldtags.html?utm_source=chatgpt.com) |
| IEEE Xplore | ("All Metadata":(imbalanc* OR "minority class" OR "skewed dataset*")) AND ("All Metadata":(oversampl* OR undersampl* OR SMOTE OR "synthetic data" OR "cost‑sensitive" OR weighting)) AND ("All Metadata":(clinic* OR medic* OR health* OR patient* OR diagnosis OR prognos*)) AND ("All Metadata":(predict* OR classification OR "logistic regression" OR "machine learning" OR "artificial intelligence")) AND (Publication Year:2009‑2024) [YouTube](https://www.youtube.com/watch?v=G3m4qDL4CME&utm_source=chatgpt.com) |
